# Large-scale study of antibody titer decay following BNT162b2 mRNA vaccine or SARS-CoV-2 infection

**DOI:** 10.1101/2021.08.19.21262111

**Authors:** Ariel Israel, Yotam Shenhar, Ilan Green, Eugene Merzon, Avivit Golan-Cohen, Alejandro A Schäffer, Eytan Ruppin, Shlomo Vinker, Eli Magen

## Abstract

**Background:** Immune protection following either vaccination or infection with SARS-CoV-2 decreases over time.

**Objective:** To determine the kinetics of SARS-CoV-2 IgG antibodies following administration of two doses of BNT162b2 vaccine, or SARS-CoV-2 infection in unvaccinated individuals.

**Methods:** Antibody titers were measured between January 31, 2021, and July 31, 2021 in two mutually exclusive groups: i) vaccinated individuals who received two doses of BNT162b2 vaccine and had no history of previous infection with COVID-19 and ii) SARS-CoV-2 convalescents who had not received the vaccine.

**Results:** A total of 2,653 individuals fully vaccinated by two doses of vaccine during the study period and 4,361 convalescent patients were included. Higher SARS-CoV-2 IgG antibody titers were observed in vaccinated individuals (median 1581 AU/mL IQR [533.8-5644.6]) after the second vaccination, than in convalescent individuals (median 355.3 AU/mL IQR [141.2-998.7]; p<0.001). In vaccinated subjects, antibody titers decreased by up to 40% each subsequent month while in convalescents they decreased by less than 5% per month. Six months after BNT162b2 vaccination 16.1% subjects had antibody levels below the seropositivity threshold of <50 AU/mL, while only 10.8% of convalescent patients were below <50 AU/mL threshold after 9 months from SARS-CoV-2 infection.

**Conclusions:** This study demonstrates individuals who received the Pfizer-BioNTech mRNA vaccine have different kinetics of antibody levels compared to patients who had been infected with the SARS-CoV-2 virus, with higher initial levels but a much faster exponential decrease in the first group.

**Funding:** This research was internally funded by Leumit Health Services (LHS) and was supported in part by the Intramural Research Program, National Institutes of Health, National Cancer Institute, Center for Cancer Research.

The content of this publication does not necessarily reflect the views or policies of the Department of Health and Human Services, nor does mention of trade names, commercial products, or organizations imply endorsement by the U.S. Government.

**Impact statement:** Large scale study display the kinetics of SARS-CoV-2 IgG antibodies present in individuals vaccinated with two doses of mRNA vaccine vs. unvaccinated patients who had recovered from the disease: initial levels of antibody are much higher in vaccinated patients, but decrease faster.

## Introduction

Immunity to severe acute respiratory syndrome coronavirus 2 (SARS-CoV-2) has been induced either through SARS-CoV-2 infection or vaccination and induces protection against reinfection or decreases the risk of clinically significant consequences(Khoury et al., 2021). While convalesced seropositive individuals have approximately 90% protection from SARS-CoV-2 reinfection, the effectiveness of vaccination has been reported as 50 - 95%(Kim et al., 2021; Lumley, 2021). Nevertheless, both the memory B cell humoral response and spike-specific CD4^+^ cellular immune responses to SARS-CoV-2 are predictably diminishing over time(Gaebler et al., 2021; Wheatley et al., 2021). Therefore, there is a great concern regarding the weakened SARS-CoV-2 immune protection both in the vaccinated and convalescent populations(Wang et al., 2021).

Israel was among the first countries to initiate a large-scale vaccination campaign, on December 20th 2020, and quickly immunized a high proportion of the adult population, achieving early control over the spread of the virus(Raz et al., 2021). More than five million Israelis (out of 9.3 million) have been fully vaccinated with two doses of the Pfizer-BioNTech vaccine as of May 26, 2021(“Covid-19 dashboard,” n.d.). However, in recent weeks, there has been a resurgence of SARS-CoV-2 cases in Israel. It is important to understand to what extent this resurgence is due to the high infectiousness of the delta variant(Chakraborty et al., 2021), lower protection of the vaccine against the delta or other variants as compared to the original strain(Faulkner et al., 2021; Martínez-Flores et al., 2021), or decreasing levels of anti-SARS-CoV-2 antibodies against all strains in vaccinated individuals(Israel et al., 2021).

Here, tracing one of these key factors, we describe the results of a large-scale study measuring the decrease rate of antibodies following administration of two doses of BNT162b2 vaccine, or SARS-CoV-2 infection in unvaccinated individuals in Israel.

## Methods

We conducted a population-based study among adult members of Leumit Health Services (LHS), a large nation-wide health maintenance organization (HMO) in Israel, which provides services to over 700,000 members. LHS has a comprehensive computerized database, continuously updated regarding subjects’ demographics, medical diagnoses, medical encounters, hospitalizations and laboratory tests. The Socio-economic status (SES) was defined according to a person’s home address. The Israeli Central Bureau of Statistics classifies all cities and settlements into 20 levels of SES. Ethnicity was also defined according to the home address of the HMO member, and categorized into three groups: General population, Ultra-Orthodox Jews and Arabs; the latter two groups are of interest because a large-scale epidemiology study showed that they had significantly higher rates of infection than the rest of the Israeli population(Muhsen et al., 2021).

All LHS members have similar health insurance coverage and similar access to healthcare services. During each physician visit, a diagnosis is entered or updated according to the International Classification of Diseases 9^th^ revision (ICD-9). The validity of chronic diagnoses in the registry has been previously examined and confirmed as high(Hamood et al., 2016; Rennert and Peterburg, 2001).

We extracted serology results and associated demographic and clinical data for members aged 18 or older, who underwent a SARS-CoV-2 serology test between January 31, 2021, and July 31, 2021, following either two vaccine injections, or documented COVID-19 infection. Patients who had had received a vaccine injection and had a documented COVID-19 infection were excluded from the study.

Baseline data from individuals included in the cohort were extracted as of May 15, 2021, including age. All the clinical diagnoses were based on ICD-9 codes. During each physician visit, a diagnosis is entered or updated according to the International Classification of Diseases 9^th^ revision (ICD-9). We tested for the main medical conditions expected to affect the severity of COVID-19 infection or the serology count in adult population: diabetes mellitus, hypertension, asthma, chronic obstructive pulmonary disease, ischemic heart disease, presence of malignancy, and chronic kidney disease.

### SARS-CoV-2 testing by real-time RT-PCR

Nasopharyngeal swabs were taken and examined for SARS-CoV-2 by real-time RT-PCR performed with internal positive and negative controls, according to World Health Organization guidelines. The Allplex 2019-nCoV assay (Seegene, Seoul, Korea) was used until March 10, 2020, after which time the COBAS SARS-Cov-2 6800/8800 assay (Roche Pharmaceuticals, Basel, Switzerland) was employed.

### SARS-CoV-2 IgG testing

Serum samples were run on the SARS-CoV-2 IgG lab-based serology blood test on the Abbot Alinity^™^ i system following the manufacturer’s instructions. In this antibody CMIA test, the SARS-CoV-2 antigen-coated paramagnetic microparticles bind to the IgG antibodies that attach to the SARS-CoV-2 spike protein (SP) in patients’ serum and plasma sample and it requires a minimum of 100 μl of serum or plasma. The resulting chemiluminescence in relative light units following the addition of anti-human IgG-labeled in comparison with the IgG II calibrator/standard indicates the strength of the response, which reflects the quantity of IgG to SP. IgG antibody levels measured by this test below 50 AU/mL are considered nonprotective. In internal testing, I Abbot AlinitI^™^ i system showed reliable results with 99.6% specificity and 100% sensitivity for COVID-19 patients tested 14 days after symptoms began (“Abbott Receives FDA Emergency Use Authorization for COVID-19 Antibody Blood Test on Alinity™ i System - May 11, 2020,” n.d.). The Abbot assay has been validated externally(Bryan et al., 2020; Perkmann et al., 2020; Zabelin et al., n.d.) with sensitivity 96.77%, specificity 99% and the caveat that these numbers depend on the boundaries of the middle range. Qualitative results and index values reported by the system were used in analyses.

The study protocol was approved by Shamir Medical Center Institutional Review Board (129-2-LEU).

### Statistical Analyses

Standard descriptive statistics were used to present the demographic characteristics of patients included in this study and their measured antibody levels. Differences in demographic and clinical characteristics between groups were analyzed using Mann-Whitney U test and Fisher’s exact test for continuous and categorical variables, respectively. Categorical data are shown in counts and percentages. Data on continuous variables are presented as mean and standard deviation, non-normal variables are displayed as median and interquartile range. Linear regression models were fit to quantify the association between time since the second vaccination in vaccinated individuals or time since the first positive PCR in convalescents, and the logarithm of antibody levels. When converted to the logarithmic scale, zero values were replaced by one. For convenience, regression coefficients are displayed in figures and tables in the natural scale (after exponentiation). Multivariable regression models were also fit to measure the effect associated for each different age categories (18-59 years, more than 60), sex, ethnic group, SES, comorbidity factor, and disease severity (presence of symptoms and admission to hospital during disease in convalescent patients).

### Software

All statistical analyses were conducted using R software version 4.0.3 (R Foundation).

## Results

During the study period, serology assays to quantify SARS-CoV-2 levels were performed for 2,653 vaccinated individuals who never had a positive SARS-Cov2 PCR test or serology test in the past, and 4,361 patients recovering from SARS-CoV-2 and who had not been vaccinated, at various times after the vaccination or infection.

Table 1 describes the demographic characteristics and serology results for tested individuals in the vaccinated population and the COVID-19 convalescent individuals, according to the time that has elapsed until the serology test. The convalescent population was younger (41.99 ± 16.09 years) than the vaccinated population (56.45 ± 15.87 years) and was characterized by lower socioeconomic status (SES) and a higher proportion of Ultra-orthodox and Arab subjects (Table 1). The mean period since the SARS-CoV-2 IgG lab-based serology test after the 2nd dose of vaccination was 101 ± 66 days, while since the first positive PCR in convalescents was 151 ± 82 days.

**Table 1.** **The demographic characteristics of tested individuals in the vaccinated and convalescent population**

|  |  | **Vaccinated** | **Convalescent** |
| --- | --- | --- | --- |
| **N** |  | 2,653 | 4,361 |
| **Age (in years)** | **mean (SD)** | 56.45 (15.87) | 41.99 (16.09) |
| **Age group**<br>**n (%)** | **18-59 years** | 1,296 (48.9%) | 3,663 (84.0%) |
|  | **≥ 60 years** | 1,357 (51.1%) | 698 (16.0%) |
| **Sex, n (%)** | **Female** | 1,604 (60.5 %) | 2,728 (62.6 %) |
|  | **Male** | 1,049 (39.5 %) | 1,633 (37.4 %) |
| **Ethnic group, n**<br>**(%)** | **Arab** | 248 (10.9 %) | 615 (14.1 %) |
|  | **General (mostly**<br>**Jewish)** | 1,633 (71.9 %) | 1,959 (44.9 %) |
|  | **Jewish Ultra-orthodox** | 389 (17.1 %) | 1,787 (41.0 %) |
| **SES, mean (SD)** |  | 9.88 (3.70) | 7.57 (3.55) |
|  | **\*missing\*** | 179 (7.24 %) | 253 (6.16 %) |
| **Body mass index**<br>**(BMI)** | **mean (SD)** | 27.79 (5.26) | 27.20 (5.74) |
|  | **\*missing\*** | 35 (1.34 %) | 92 (2.16 %) |
| **BMI category,**<br>**n (%)** | **< 18.5 Underweight** | 46 (1.8 %) | 135 (3.2 %) |
|  | **18.5 - 25 Normal** | 780 (29.8 %) | 1,468 (34.4 %) |
|  | **25 - 30 Overweight** | 994 (38.0 %) | 1,470 (34.5 %) |
|  | **30 - 35 Obese I** | 550 (21.0 %) | 781 (18.3 %) |
|  | **35 - 40 Obese II** | 195 (7.5 %) | 304 (7.1 %) |
|  | **40 - Obese III** | 51 (1.9 %) | 105 (2.5 %) |
|  | **\*missing\*** | 37 (1.4 %) | 98 (2.2 %) |
| **co-morbidities,**<br>**n (%)** | **diabetes mellitus** | 659 (24.8 %) | 493 (11.3 %) |
|  | **hypertension** | 1,140 (43.0 %) | 808 (18.5 %) |
|  | **asthma** | 299 (11.3 %) | 409 (9.4 %) |
|  | **COPD** | 265 (10.0 %) | 137 (3.1 %) |
|  | **ischemic heart disease** | 325 (12.3 %) | 183 (4.2 %) |
|  | **solid tumor** | 342 (12.9 %) | 172 (3.9 %) |
|  | **chronic renal disease** | 199 (7.5 %) | 58 (1.3 %) |
| **Time (in days)**<br>**since... mean (SD)** | **2<sup>nd</sup> vaccination** | 101.35 (65.73) | - |
|  | **first positive PCR** | - | 151.17 (82.32) |
| **Time (in days)**<br>**since vaccination**<br>**or positive PCR**<br>**by 30 days**<br>**intervals,**<br>**n (%)** | **0-29** | 556 (21.0 %) | 269 (6.2 %) |
|  | **30-59** | 456 (17.2 %) | 499 (11.4 %) |
|  | **60-89** | 289 (10.9 %) | 341 (7.8 %) |
|  | **90-119** | 200 (7.5 %) | 331 (7.6 %) |
|  | **120-149** | 170 (6.4 %) | 700 (16.1 %) |
|  | **150-179** | 542 (20.4 %) | 735 (16.9 %) |
|  | **180-209** | 440 (16.6%) | 587 (13.5 %) |
|  | **210-239** | - | 365 (8.4 %) |
|  | **240-269** | - | 161 (3.7 %) |
|  | **270-** | - | 373 (8.6 %) |

SES: socio-economic status; BMI: body mass index; COPD: chronic obstructive pulmonary disease

Table 2a and 2b display SARS-CoV-2 IgG antibody titers measured in vaccinated and convalescent individuals, in intervals spaced 30 days apart, since second vaccination (for the vaccinated) or first positive PCR (for convalescents). Lab-based serology is available for up to six months following vaccination for the vaccinated and up to nine months for convalescent patients. The age distribution of the patients for which serology was tested varies slightly throughout the follow-up, so it is indicated in the tables.

**Table 2a.** Serology results of vaccinated individuals by 30 days intervals since second vaccination.

| Time since second vaccine injection (in days) |  | 0-29 | 30-59 | 60-89 | 90-119 | 120-149 | 150-179 | 180- |
| --- | --- | --- | --- | --- | --- | --- | --- | --- |
| N |  | 556 | 456 | 289 | 200 | 170 | 542 | 440 |
| Age (in years) |  | 53.76 (16.90) | 55.41 (16.66) | 55.64 (16.52) | 53.31 (16.75) | 53.29 (16.29) | 56.34 (13.76) | 64.23 (12.30) |
| Sex, n (%) | Female | 318 (57.2 %) | 273 (59.9 %) | 173 (59.9 %) | 129 (64.5 %) | 104 (61.2 %) | 358 (66.1 %) | 249 (56.6 %) |
|  | Male | 238 (42.8 %) | 183 (40.1 %) | 116 (40.1 %) | 71 (35.5 %) | 66 (38.8 %) | 184 (33.9 %) | 191 (43.4 %) |
| SARS-CoV-2 IgG antibody level | mean | **12153** | **6848** | **3476** | **2383** | **1552** | **1122** | **765** |
|  | (SD) | **(9875)** | **(6340)** | **(4582)** | **(3266)** | **(2103)** | **(1431)** | **(948)** |
|  | median [IQR] | **9913**<br>[3650-18733] | **5106**<br>[2109-9601] | **2159**<br>[1039-4169] | **1323**<br>[549-3126] | **1071**<br>[471-1901] | **764**<br>[385-1343] | **447**<br>[205-966] |

**Table 2b.** Serology results of convalescent patients by 30 days intervals since second first positive PCR test.

| Time since first positive PCR (in days) |  | 0-29 | 30-59 | 60-89 | 90-119 | 120-149 | 150-179 | 180-209 | 210-239 | 240-269 | 270- |
| --- | --- | --- | --- | --- | --- | --- | --- | --- | --- | --- | --- |
| N |  | 269 | 499 | 341 | 331 | 700 | 735 | 587 | 365 | 161 | 373 |
| Age (in years) |  | 39.96 (14.78) | 41.10 (14.95) | 45.07 (16.53) | 43.52 (16.32) | 42.90 (16.23) | 40.95 (16.13) | 41.39 (16.14) | 40.59 (15.32) | 41.75 (17.42) | 43.18 (17.10) |
| Sex, n (%) | Female | 157 (58.4 %) | 326 (65.3 %) | 234 (68.6 %) | 219 (66.2 %) | 400 (57.1 %) | 461 (62.7 %) | 371 (63.2 %) | 230 (63.0 %) | 112 (69.6 %) | 218 (58.4 %) |
|  | Male | 112 (41.6 %) | 173 (34.7 %) | 107 (31.4 %) | 112 (33.8 %) | 300 (42.9 %) | 274 (37.3 %) | 216 (36.8 %) | 135 (37.0 %) | 49 (30.4 %) | 155 (41.6 %) |
| SARS-CoV-2 IgG antibody level | mean | **1914** | **1739** | **1552** | **1195** | **1079** | **860** | **904** | **850** | **901** | **731** |
|  | (SD) | **(3870)** | **(2972)** | **(2522)** | **(2406)** | **(2556)** | **(1962)** | **(2384)** | **(2104)** | **(1739)** | **(1280)** |
|  | median [IQR] | **490**<br>[109-1869] | **586**<br>[212-1908] | **538**<br>[247-1723] | **377**<br>[165-1080] | **329**<br>[140-886] | **312**<br>[138-8301] | **278**<br>[125-751] | **278**<br>[105-727] | **351**<br>[124-919] | **314**<br>[116-783] |

We observe considerably higher titers in the first month following the second vaccination (median 9913, IQR [3650-18,733]) than in convalescent patients after SARS-CoV-2 infection (median 490, IQR [109-1869] in the first month). In the convalescent subjects, the maximal mean antibody response was observed at 3 months after the documented COVID-19 infection, then the mean SARS-CoV-2 IgG antibody titer decreases slightly each subsequent month from the highest mean antibody response. The vaccination with the BNT162b2 vaccine-elicited much higher antibody titers at 3 months compared to the titers collected in serum from convalescent patients. However, in these vaccinated individuals who never had a positive PCR test, the mean SARS-CoV-2 IgG antibody titer decreased by up to 40% each subsequent month from the highest mean antibody response. Consequently, we observed in BNT162b2 vaccinated subjects a worrisome decline in the proportion of people whose antibody levels are below the seropositivity threshold of <50 AU/mL (considered non-protective) from 5.8% in the first 3 months, to 16.1% after 6 months (**Figure 1a**) while only 10.8% of convalescent patients are below the 50AU/mL threshold after 9 months (**Figure 1b**).

**Figure 1a.**
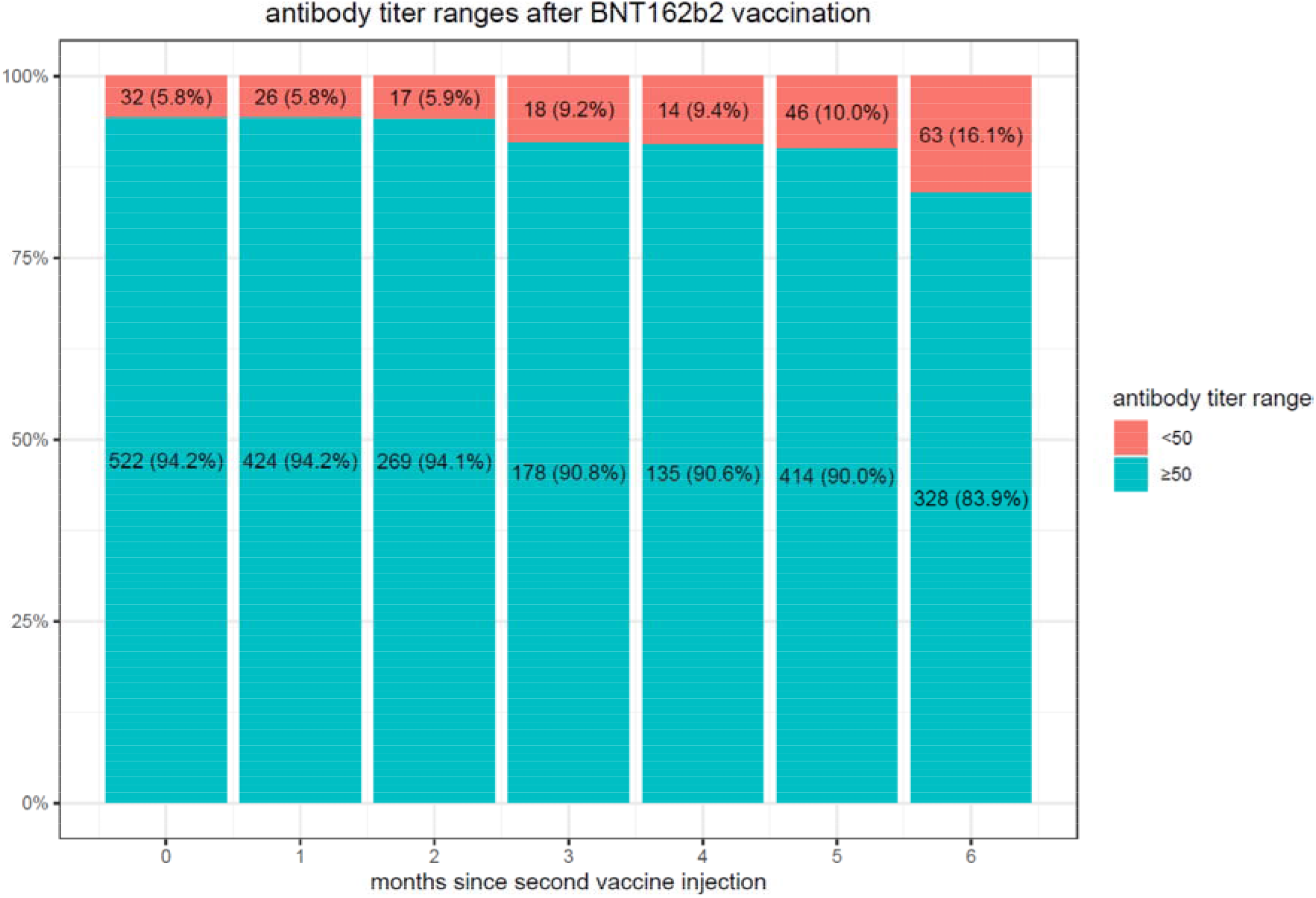

**Figure 1b.**
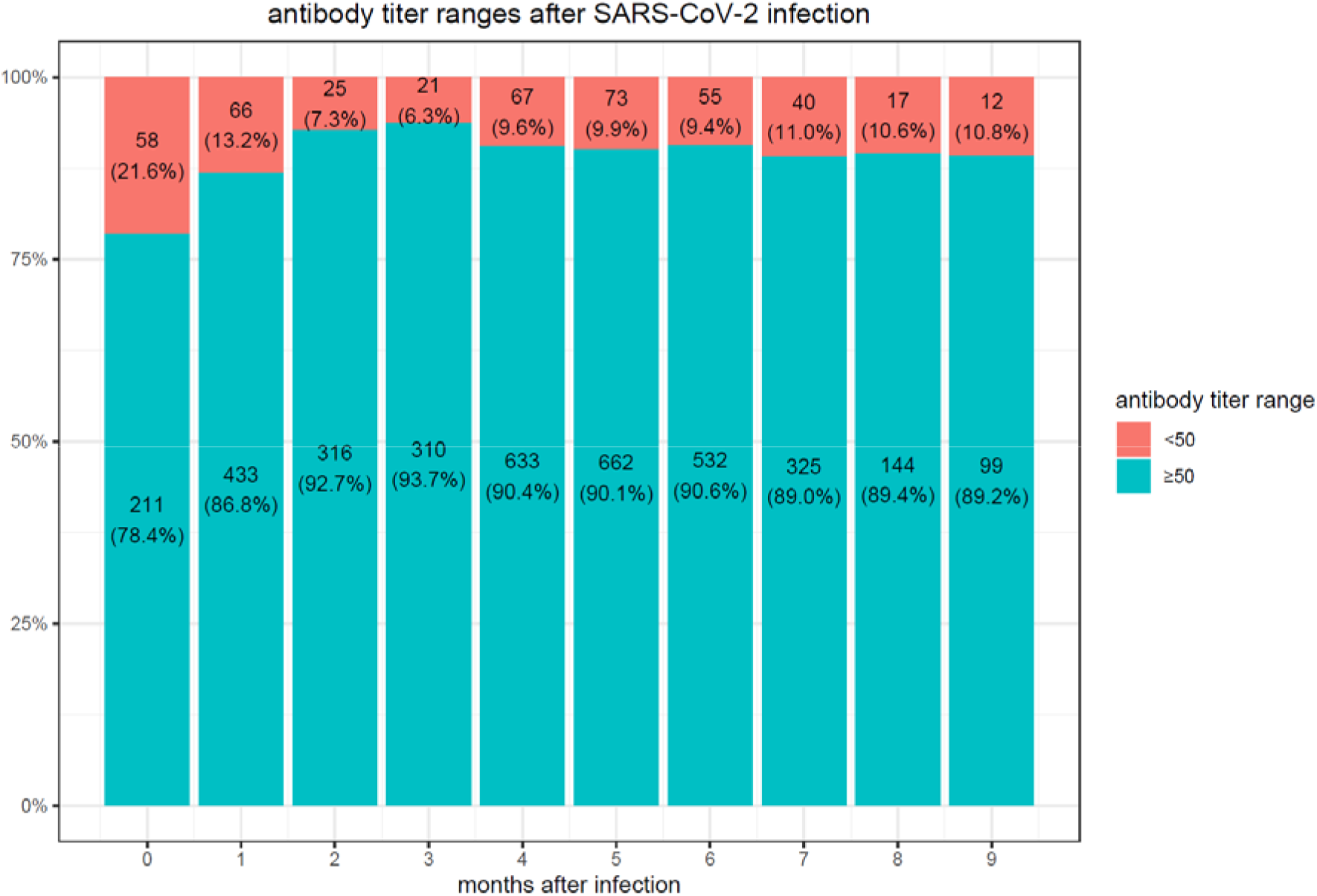

**Figure 2a.**
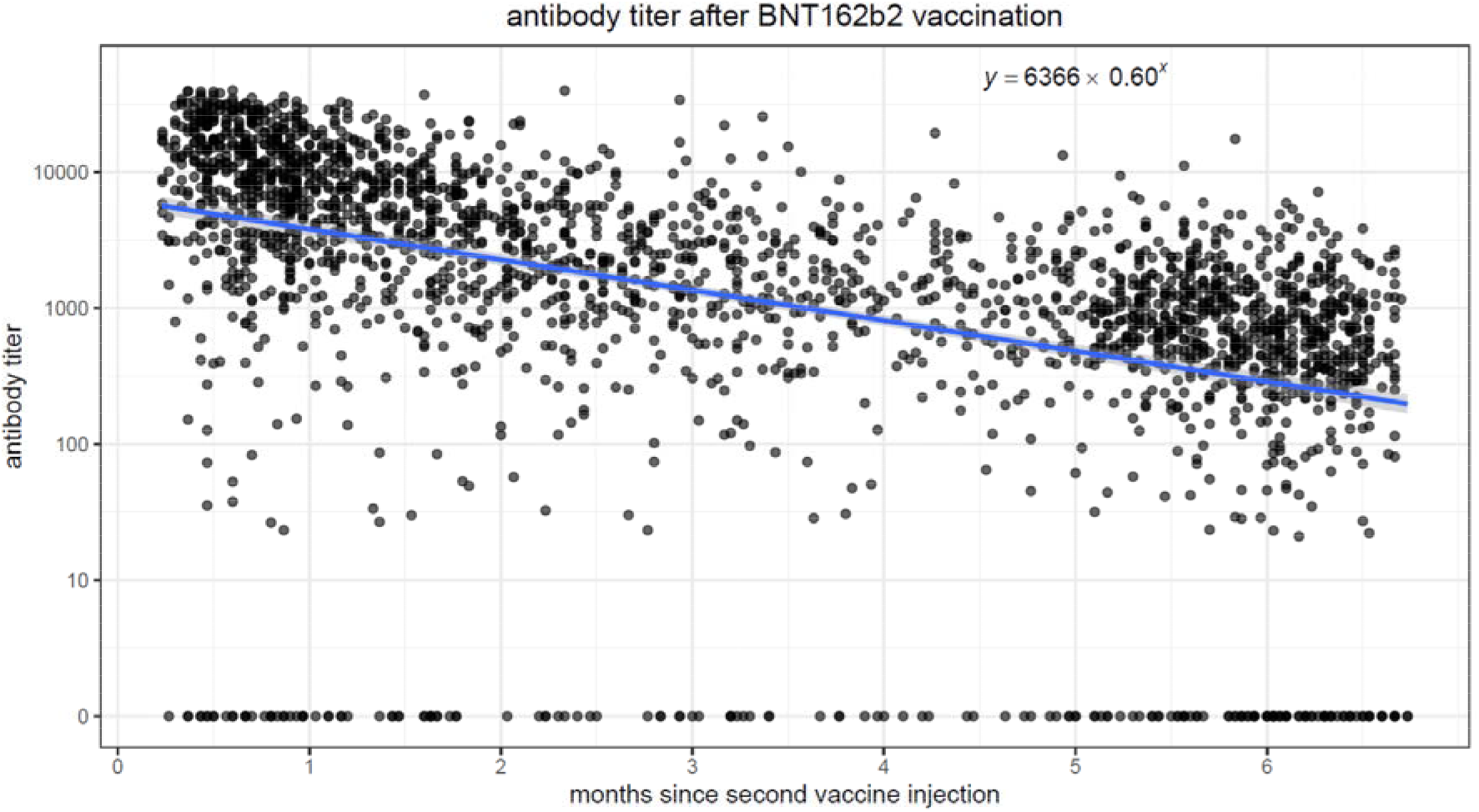

**Figure 2b.**
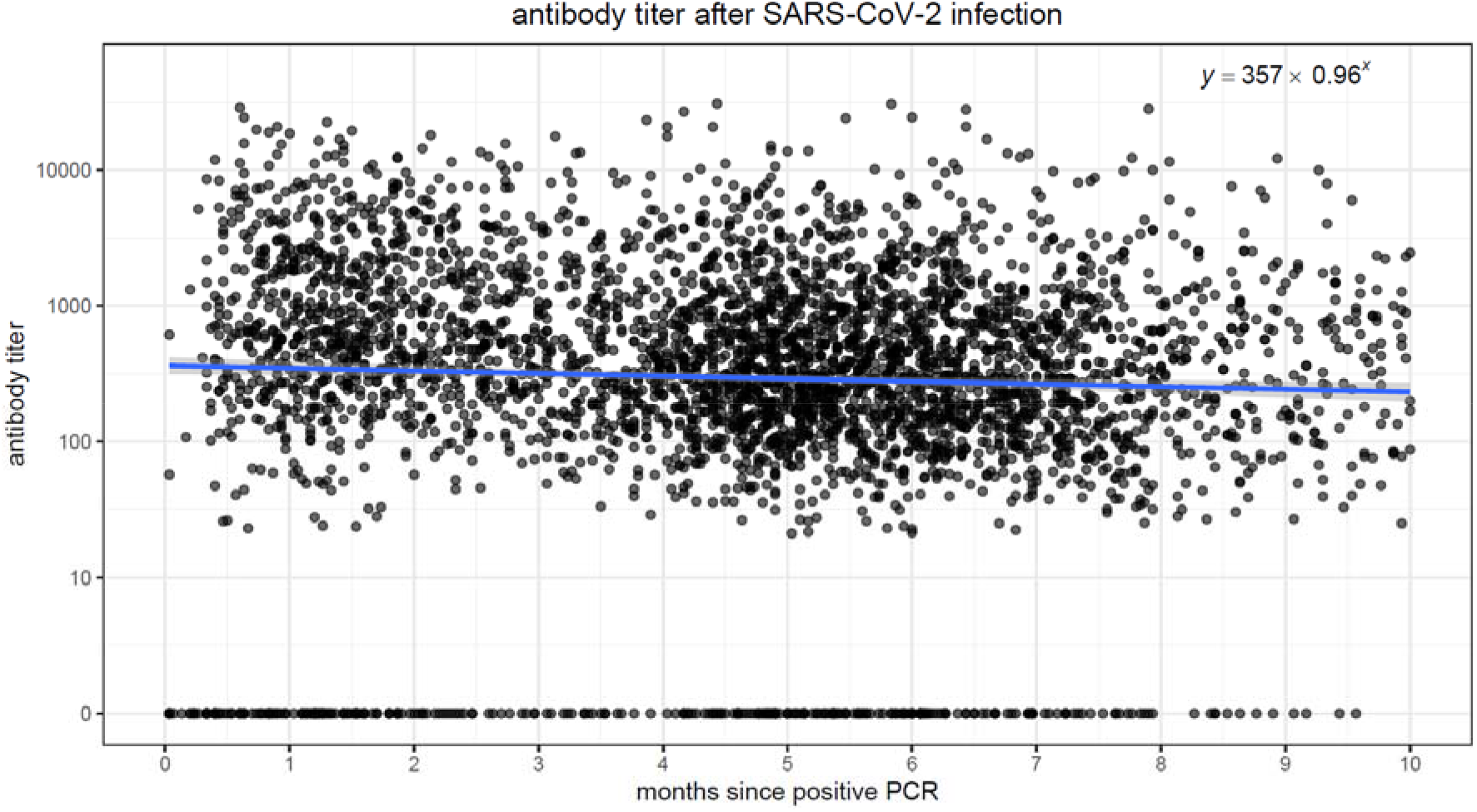

We performed linear regression models to quantify the association between elapsed time and antibody levels, in both vaccinated and convalescent individuals. In both populations, there was a strong association (p<0.001) between elapsing time and antibody titers. **Figure 1a. and 1b**. display scatter plots with antibody titers plotted against elapsed time. In the vaccinated population, we observe higher initial antibody titers (intercept of 6366 at time zero), but the titers quickly drop, decreasing by 40% each passing month. Conversely, in the convalescent population, initial titers are lower (intercept of 357 at time zero), but the titers decrease much more slowly, by ~4% every month.

To assess possible effects of age, sex, ethnic group, SES in addition to time since the second vaccination or since the first positive PCR in convalescents on antibody levels, we performed multivariable regression models on the vaccinated and convalescent cohorts (Table 3). In both populations, there was a strong association (p<0.001) between elapsed time and antibody titers: each month was associated with a mean decay factor of 0.623 [95% CI 0.599-0.649] in vaccinated patient, while for convalescent patients the decrease was only by a factor of 0.960 [95% CI 0.939-0.982]. Among the vaccinated, antibody titers decreased with older age (factor 0.790 [95% CI 0.644-0.969] for age≥60), chronic renal disease (factor 0.200 [95% CI 0.143-0.281]), underweight (factor 0.359 [95% CI 0.144-0.893] for BMI<18.5), solid malignancy (factor 0.642 [95% CI 0.494-0.834]), COPD (factor 0.643 [95% CI 0.479-0.863]), patients with diabetes mellitus (factor 0.720 [95% CI 0.579-0.894]), and hypertension (factor 0.786 [95% CI 0.639-0.966]); they were increased in females (factor 1.243 [95% CI 1.035-1.492]) and in Arab and Jewish Ultra-orthodox subjects. In the convalescent, antibody titers were higher for symptomatic patients (factor 1.811 [95% CI 1.531-2.142]), those who had been admitted to the hospital (factor 3.323 [95% CI 2.217-4.980]) and those with risk factors for severe disease: older age (factor 1.546 [95% CI 1.269-1.884] for age≥60), obesity (factor 1.839 [95% CI 1.166-2.899] for BMI>35), diabetes mellitus (factor 1.354 [95% CI 1.093-1.678]), hypertension (factor 1.254 [95% CI 1.036-1.518]), and chronic renal disease (factor 1.965 [95% CI 1.134-3.407]).

**Table 3.**
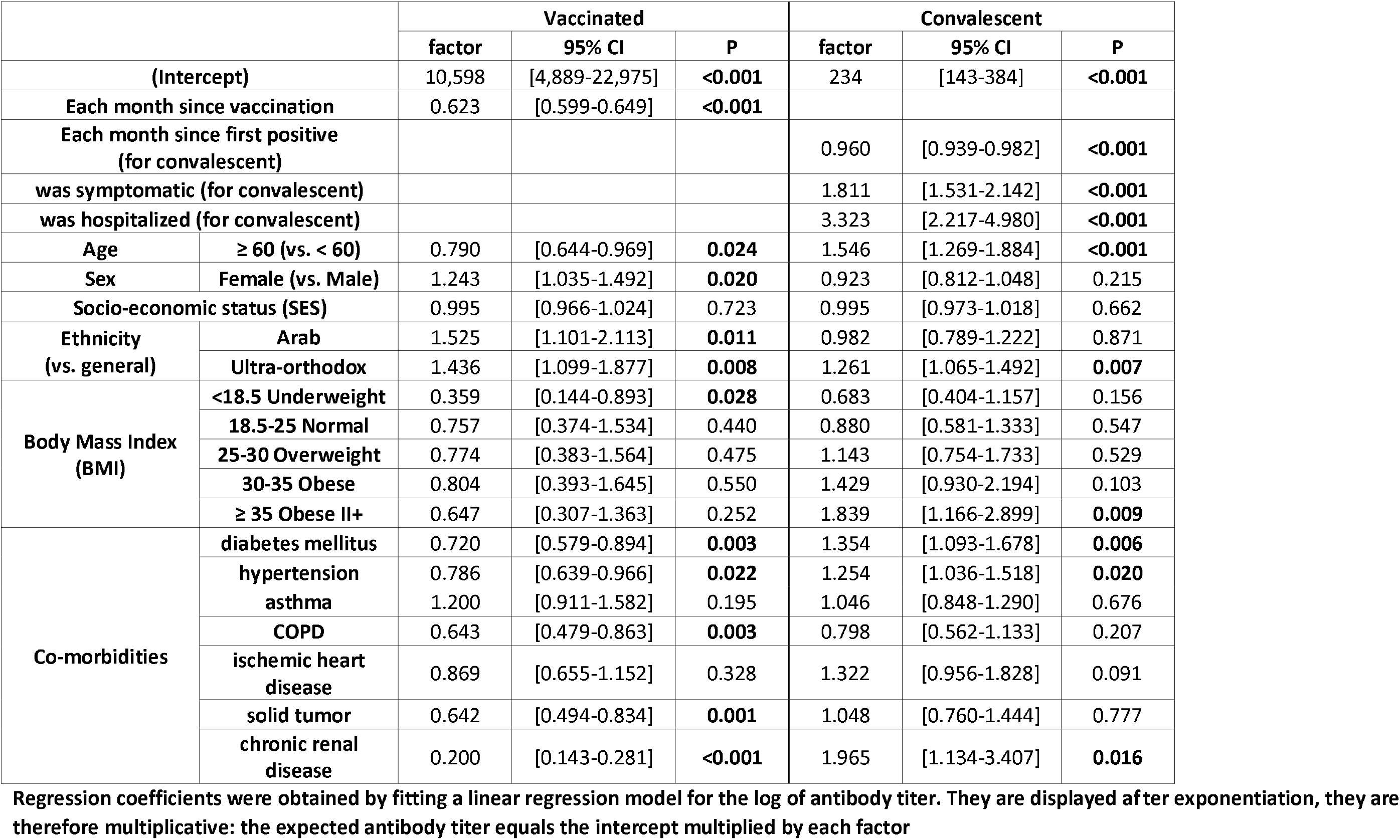
Linear regression models of SARS-CoV-2 IgG antibody titer.

## Discussion

In this large population of individuals tested for SARS-CoV-2 antibody titer following either vaccination or documented COVID-19 infection, we correlated antibody titers to the time that has elapsed since exposure to vaccine or virus. Among never infected individuals who received the Pfizer-BioNTech mRNA vaccine, we found higher initial antibody levels followed by a faster decrease rate compared to patients who had been infected with the SARS-CoV-2 virus. As a corollary of that, the proportion of vaccinated individuals whose antibody levels drop below the threshold thought to be protective is increasing substantially by the fifth month, while it is uncommon in convalescent individuals.

Real world data about the time evolution of SARS-CoV-2 antibodies after vaccinations against SARS-CoV-2 and after the COVID-19 infection are limited. Several studies reported on the humoral response following the BNT162b2 mRNA COVID-19 vaccination and found that SARS-CoV-2 antibody titers 4 to 6 months after infection decline more slowly than in the initial months after the infection (Ebinger et al., 2021; Padoan et al., 2021; Tré-Hardy et al., 2021). At least two recent studies reported that antibodies could be detected up to 11 months after infection and provided evidence that that these antibodies originated in memory B cells, not plasmablasts(Tong et al., 2021; Turner et al., 2021a). Generation of SARS-CoV-2 memory B cells(De Gasparo et al., 2021; Schoof et al., 2020; Xu et al., 2021) is likely necessary for long-term protection since that is the mechanism by which most anti-viral vaccines work(Iwasaki, 2016).

The strength of our study is that it provides such information in a relatively large cohort of both vaccinated individuals and patients recovering from SARS-CoV-2. It shows that the declining slope of antibodies in vaccinated individuals is much steeper than in convalescent individuals. Other studies reported on the persistence of the humoral response in vaccinated subjects, but the follow-up of the participants was usually below 3 months(Salvagno et al., 2021; Tré-Hardy et al., 2021; Turner et al., 2021b). This study has several limitations. First, given the observational design, there is potential for unmeasured confounding factors. In particular, participants in this study were individuals who elected to have a serology test for SARS-CoV-2 during the study period, many of them as part of a survey. Individuals may have variable reasons for accepting or refusing the offered serology test, which may have affected the results of this survey. Additionally, even though we adjusted our results for known factors that could affect antibody production and decay, additional factors may have affected the results.

The BNT162b2 mRNA COVID-19 vaccine has been shown to stimulate production of antibodies to several SARS-CoV-2 proteins, not just the spike protein, suggesting that the vaccine provides short-term protection against variant strains(Amanat et al., 2021; Turner et al., 2021b). Combining these results with ours suggests that the recent surge in breakthrough infections in fully vaccinated individuals is due at least in part to declining levels of antibodies and not solely due to the variant strains of SARS-CoV-2. In a recent single-center, prospective, cross-sectional cohort study, children’s immunity was found to decline 4 months after the COVID-19 infection(Breuer et al., 2021). Lyer et al. reported that anti-S-protein antibodies reserved neutralizing abilities and persevered for up to 75 days after SARS-CoV-2 infection in >95% of patients(Iyer et al., 2020). Gudbjartsson et al. showed that SARS-CoV-2 IgG levels do not wane up to 4 months after infection(Gudbjartsson et al., 2020).

Shortly after vaccination, the BNT162b2 mRNA vaccine was shown to protect against COVID-19 through multiple beneficial mechanisms, by eliciting robust CD4^+^ and CD8^+^ T cell responses and strong antibody responses to the receptor-binding domain (RBD) of the SARS-CoV-2 spike protein, with RBD-binding IgG concentrations clearly above those seen in serum from a cohort of individuals who had recovered from COVID-19(Sahin et al., 2020). These effective humoral and cellular immune responses were observed a week after the booster dose, with the negligible immune response between the first and second doses(Sahin et al., 2020).

Bone marrow plasma cells (BMPCs) are an essential source of medium-term protective antibodies both after vaccination and infection(Halliley et al., 2015; Nutt et al., 2015), but longer-term protection likely requires memory B cells(Iwasaki, 2016; Turner et al., 2021a). Individuals who have recovered from COVID-19 have a significantly lower risk of SARS-CoV-2 reinfection(Hall et al., 2021). A recent 12-month longitudinal study was published with 1,782 plasma samples from 869 convalescent plasma donors in Wuhan, China. This study has shown that among COVID-19 plasma donors the positive rate of IgG antibody against the SARS-CoV-2 receptor binding domain (RBD) in the spike protein exceeded 70% for 12 months post-diagnosis(C. Li et al., 2021). In our study, we show that following vaccination, the levels of anti-SARS-CoV-2 antibodies decrease rapidly, indicating that BMPCs may not be created adequately and therefore anti-SARS-CoV-2 humoral immunity might be transient(Ibarrondo et al., 2020; Seow et al., 2020). After infection, SARS-CoV-2 proteins and nucleic acids could remain in the gut for at least two months, boosting the continued antibody evolution in germinal centers, preferring epitopes overlapping with the ACE2-binding site on the RBD(Gaebler et al., 2021).

It is widely accepted that neutralizing serum antibodies provide a strong protective role from SARS-CoV-2 in both nonhuman primates animal models and in humans(D. Li et al., 2021; McMahan et al., 2020). The assay used in our study does not specially measure neutralizing antibody, nevertheless, a high correlation was observed between a surrogate virus neutralization assay and other assays such as Roche Elecsys anti-S pan-Ig assay(L’Huillier et al., 2021).

Remarkably, after BNT162b2 mRNA vaccination, we observed higher SARS-CoV-2 antibody titers in the convalescent individuals aged ≥ 60 years, while in the vaccinated population higher SARS-CoV-2 antibody titers were seen in younger patients. Clinically, in a recent study performed in our health organization among individuals who had received two doses of the BNT162b2 vaccine, we observed a significantly higher rate of SARS-CoV-2 infection among patients who have received their second vaccine dose for more than 146 days: the increase was significant for all age groups, with the strongest increase observed for patients aged 60 or more(Israel et al., 2021). The decrease of SARS-CoV-2 IgG antibodies observed in the present study provides one explanation for the increased infection rate found after 146 days in this age group and may warrant a third vaccine dose before the fifth month in populations at risk. Our observations call for replication in other populations to further correlate of protection against SARS-CoV-2 reinfection and/or COVID-19 disease and the duration of antibody-mediated protection.

## Data Availability

Data were obtained from patients' electronic health records, and IRB approval restrains its use to researchers inside Leumit Health Services.

## Contributors

All authors provided final approval to publish The corresponding author attests that all listed authors meet authorship criteria and that no others meeting the criteria have been omitted. AI is the guarantor.

## Data Availability Statement

This study is based on real-world patient data, including demographics, comorbidity factors, that cannot be communicated due to patient privacy concerns.

## Abbreviations

COVID-19: Coronavirus disease of 2019
SARS-CoV-2: Severe acute respiratory syndrome coronavirus 2
mRNA: Messenger RNA
LHS: Leumit Health Services
HMO: Health maintenance organization
SES: Socio-economic status
BMI: Body mass Index
IQR: Interquartile range
RT-PCR: Reverse transcription polymerase chain reaction
IgG: Immunoglobulin G
BMPCs: Bone marrow plasma cells
ACE2: Angiotensin-converting enzyme 2

## Acknowledgements

The content of this publication does not necessarily reflect the views or policies of the Department of Health and Human Services, nor does mention of trade names, commercial products, or organizations imply endorsement by the US Government.

